# Detection of SARS-CoV-2 N501Y mutation by RT-PCR to identify the UK and the South African strains in the population of South Indian state of Telangana

**DOI:** 10.1101/2021.03.27.21254107

**Authors:** Safaa Muneer Ahmed, Smita Rao Juvvadi, Rakesh Kalapala, Jagadeesh Babu Sreemanthula

**Author notes:** Correspondence to Jagadeesh Babu Sreemanthula.

## Abstract

**Objective:** To detect N501Y mutation of the SARS-CoV-2 spike protein by RT-PCR to distinguish (B.1.1.7) UK and (501Y.V2) South African strains from others in the population of Telangana and to determine its clinical implications.

**Methods:** A primer-probe mix that specifically detects the mutated N501Y strain by real time RT-PCR was designed. 93 samples that were reported positive for COVID-19 by our laboratory in the month of February 2021 were tested using our own primer-probe mix for the presence of N501Y by RT-PCR. The results of RT-PCR were validated by Sanger sequencing in representative samples. Sanger sequencing of other defining spike mutations of B.1.1.7 (del 69-70, del 144, N501Y, A570D, D614G, P681H, T716I, S982A and D1118H) and 501Y.V2 (K417N, E484K, N501Y and D614G) was also investigated.

**Findings:** Out of 93 COVID-19 positive samples, 12 samples are detected positive for N501Y by RT-PCR. Sanger sequencing of these 12 samples further confirmed the presence of N501Y and other mutations that are characteristic of UK strain (B.1.1.7). The South African strain (501Y.V2) is not detected in any of our samples in this study. But, the E484K mutation that is characteristic of 501Y.V2 is detected in one N501Y negative sample.

**Conclusion:** Strain-specific RT-PCR for N501Y was developed and validated with Sanger sequencing. Such strategy facilitates quick surveillance for more transmissible and more vaccine resistant strains.

## Introduction

As the world is fighting hard to tame the Covid-19 pandemic, the virus is fighting harder to best adapt to humans and evade vaccines by mutating itself. It is necessary to study sequence variations constantly to control highly transmissible and vaccine-resistant strains. By June 2020, the Wuhan originated D614 virus made a complete global transition to G614, probably to ensure its own survival^1^.Now, it has further evolved into more transmissible UK (B.1.1.7) and potentially more neutralization resistant South African (501Y.V2) strains^2, 3, 4^. The UK variant is defined by nine spike protein mutations i.e, del 69-70, del 144, N501Y, A570D, D614G, P681H, T716I, S982A and D1118H^5^. The South African variant is defined by four spike protein mutations i.e, K417N, E484K, N501Y and D614G^3^. Another lineage (501Y.V3) has been idenfied in Manaus, Brazil defined by K417T, E484K and N501Y^6^. These strains if not monitored could result in an epidemic rebound^7^.

This study was aimed to investigate the percentage of Covid-19 positive samples that are tested in our laboratory bearing the N501Y mutation, which is common in both the UK and the South African variants. Since Sanger-sequencing of every positive sample for this mutation is both time-taking and expensive, we designed a primer-probe mix to detect specifically the N501Y mutation of the novel Coronavirus to filter out UK and South African strains from other strains by Real time-Reverse transcriptase polymerase chain reaction (RT-PCR).We further validated the results of RT-PCR by Sanger sequencing all N501Y positive and a limited number of N501Y negative samples. We also demonstrated Sanger sequencing of all defining mutations of the UK strain and E484K mutation to distinguish South African strain from the UK strain.

## Methods

Sample collection: Nasopharyngeal/Oropharyngeal Swabs collected in Viral Transport Medium which were sent to our laboratory for the detection of SARS-CoV-2 were tested using ICMR approved RT-PCR kits. Among these, 93 Viral RNA templates that tested positive for SARS-CoV-2 were randomly selected and saved.

Primer design: The oligonucleotides (Table 1) that were used in RT-PCR and Sanger sequencing were synthesized.

**Table 1.**
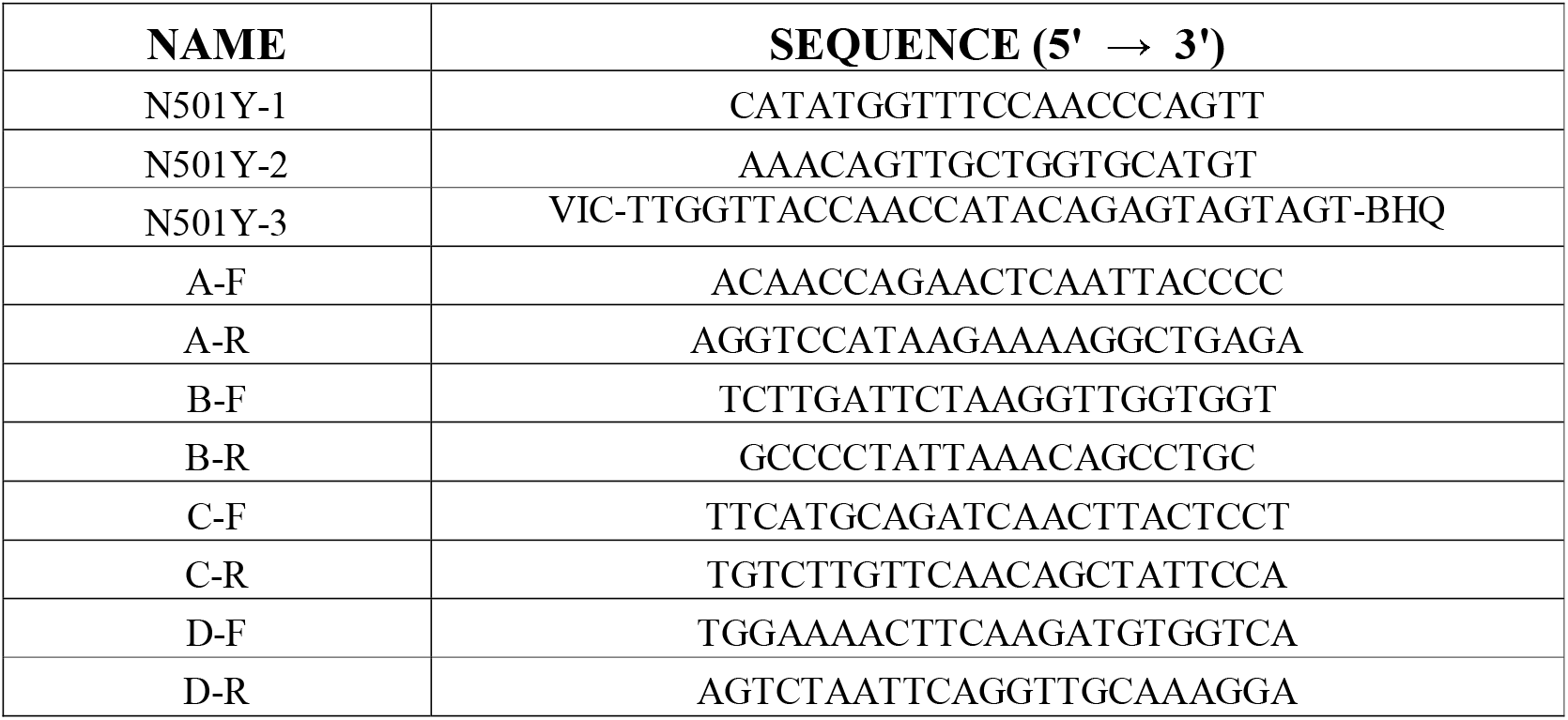
Oligonucleotide sequences used.

Real time RT-PCR: The reaction (total volume 30 µl) was set up by adding 15 µl 2X PCR mastermix (M7502, Promega), 5 µl RTase mix (M-NCOV-01, SD BIOSENSOR), 0.5 µl 50X Rox Reference Dye LSR (ST0290, TAKARA), 1µl forward primer N501Y-1 (10 pmol/µl), 1µl reverse primer N501Y-2 (10 pmol/µl), 1µl VIC probe N501Y-3 (10 pmol/µl) and 6.5 µl RNA template. The thermalcycler (QuantStudio 5 System) conditions were reverse transcription at 50°C for 20 min; initial denaturation at 95°C for 5 min; followed by 50 cycles of denaturation at 95°C for 15 sec, annealing at 55°C for 15sec and extension at 72°C for 15 sec with data collection; and final extension at 72°C for 15 sec with data collection. The experiment type was selected as Presence/Absence and the Chemistry was selected as TaqMan Reagents. The Passive reference dye was chosen as ROX. The Reporter was selected as VIC and the Quencher as NFQ-MGB. A total of 96 samples were included in the run that includes 93 test samples, a known N501Y positive control (PC), a known N501Y negative control (NC) and a No Template Control (NTC).

Sanger sequencing: The PCR reaction (total volume 25 µl) was set up by adding 12.5 µl 2X PCR mastermix (M7502, Promega), 5 µl RTase mix (M-NCOV-01, SD BIOSENSOR), 1µl forward primer, 1 µl reverse primer and 5.5µl RNA template. The thermal conditions were reverse transcription at 50°C for 20 min, followed by 40 cycles of denaturation at 95°C for 45 sec, annealing at 55°C for 45 sec, extension at 72°C for 45 sec and final extension at 72°C for 5 min. The primer pair A-F and A-R amplifies a 482 bp product that covers del 69-70 and del 144 mutations. The primer pair B-F and B-R amplifies a 639 bp product that covers E484K, N501Y, A570D and D614G mutations. The primer pair C-F and C-R amplifies a 456 bp product that covers P681H and T716I mutations. The primer pair D-F and D-R amplifies a 605 bp product that covers S982A and D1118H mutations. The amplified products (5 µl) were electrophoresed in 2% agarose gel and subjected to Sanger sequencing on Applied Biosystems SeqStudio Genetic Analyzer by following manufacturer’s instructions.

## Results

Real time RT-PCR: The amplification plot type was chosen as ΔRn vs Cycle and graph type as Linear. The Analysis settings were Baseline Start 4, Baseline End 22 and the Threshold 0.0125. The amplification plot was analysed for samples with threshold above 0.0125. A total of 12 samples exceeded the cut-off and were recorded as N501Y positive (Figure 1A). The amplification plot for Controls PC, NC and NTC was also recorded (Figure 1B).

**Figure 1.**
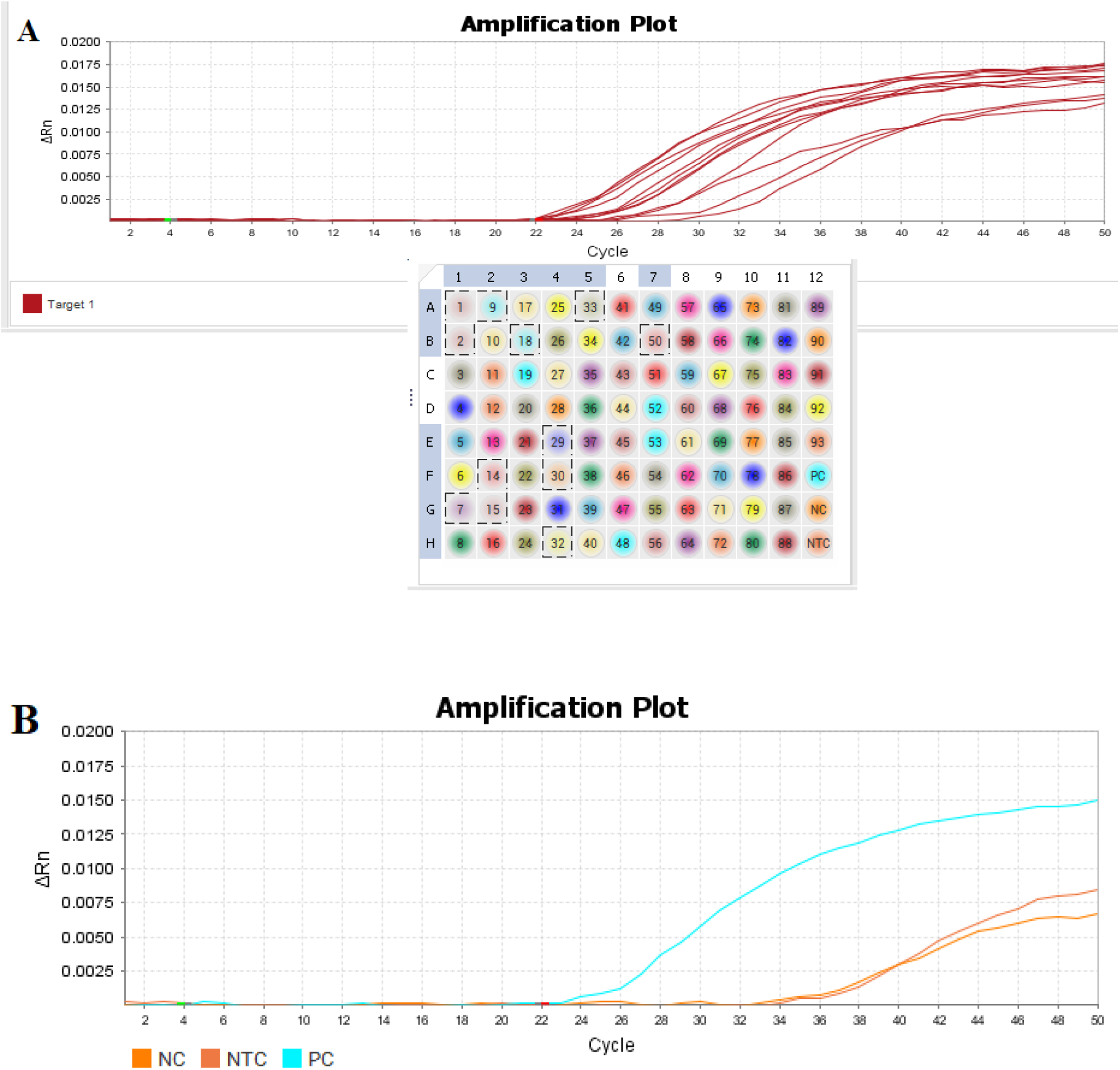
(A) Amplification plot for samples above threshold 0.0125. Out of 93, 12 samples passed the cut-off and recorded as N501Y positive. (B) Amplification plot for Control samples PC (Known N501Y positive), NC (Known N501Y negative) and NTC (No Template Control).

Sanger sequencing: The presence and absence of N501Y as detected by RT-PCR was also confirmed by Sanger sequencing. The data was analysed by CodonCode Aligner software. The chromatogram showing Y501 was presented, the un-mutated counterpart N501 provided for comparison (Figure 2).None of the twelve N501Y positive samples had E484K mutation suggesting the absence of South African strain. The chromatogram showing deletion 69-70 was presented, the un-deleted counterpart provided for comparison (Figure 3). The chromatogram showing deletion 144 was presented, the un-deleted counterpart provided for comparison (Figure 4). The chromatogram showing D570 was presented, the un-mutated counterpart A570 provided for comparison (Figure 5). The chromatogram showing G614 was presented, the un-mutated counterpart D614 is no longer available (Figure 6). The chromatogram showing H681 was presented, the un-mutated counterpart P681 provided for comparison (Figure 7). The chromatogram showing I716 was presented, the un-mutated counterpart T716 provided for comparison (Figure 8). The chromatogram showing A982 was presented, the un-mutated counterpart S982 provided for comparison (Figure 9). The chromatogram showing H1118 was presented, the un-mutated counterpart D1118 provided for comparison (Figure 10). The chromatogram showing K484 that was detected in N501Y negative sample was presented, the counterpart E484 provided for comparison (Figure 11).

**Figure 2.**
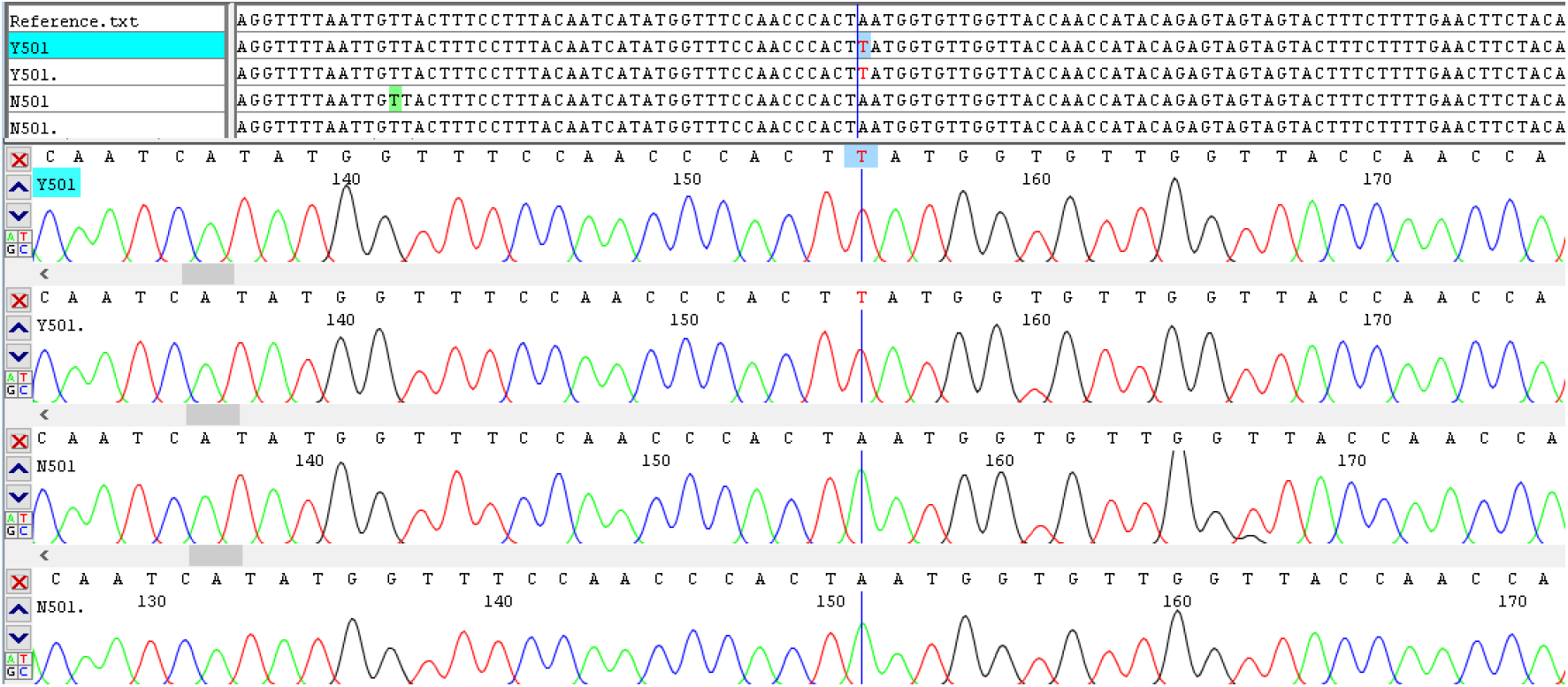
Chromatogram showing N501Y (Asn501Tyr) mutation of SARS-CoV-2 spike protein. The AAT>TAT conversion at codon 501 is highlighted.

**Figure 3.**
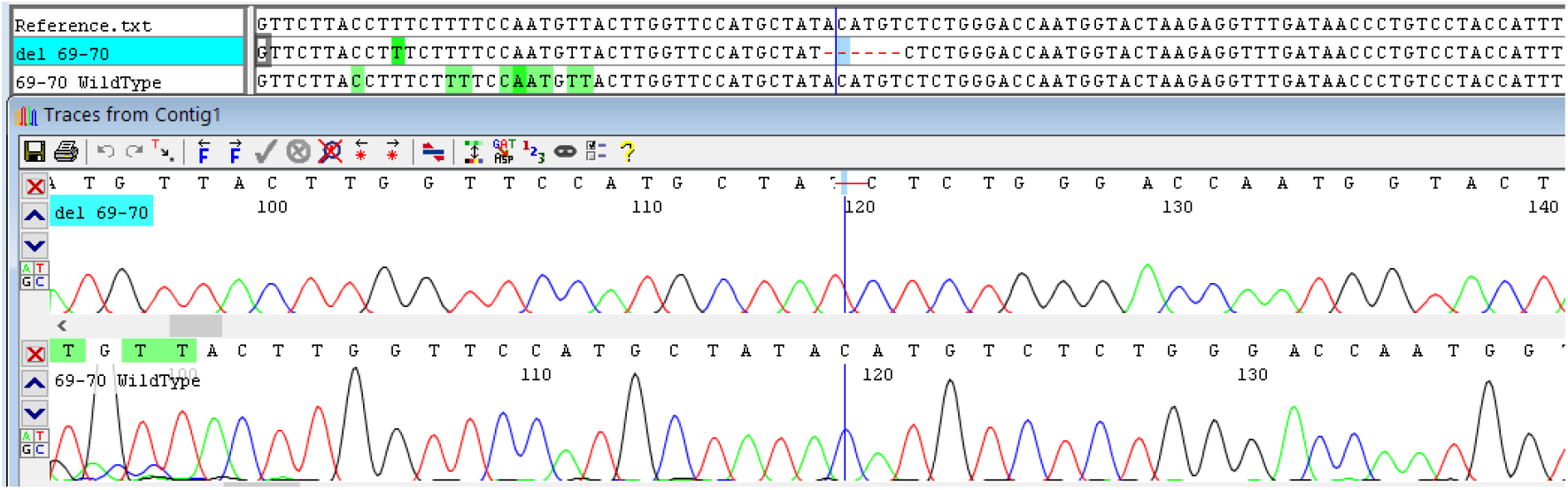
Chromatogram showing del 69-70 mutation of SARS-CoV-2 spike protein. The WildType chromatogram without codon 69 and 70 deletion is shown for comparison.

**Figure 4.**
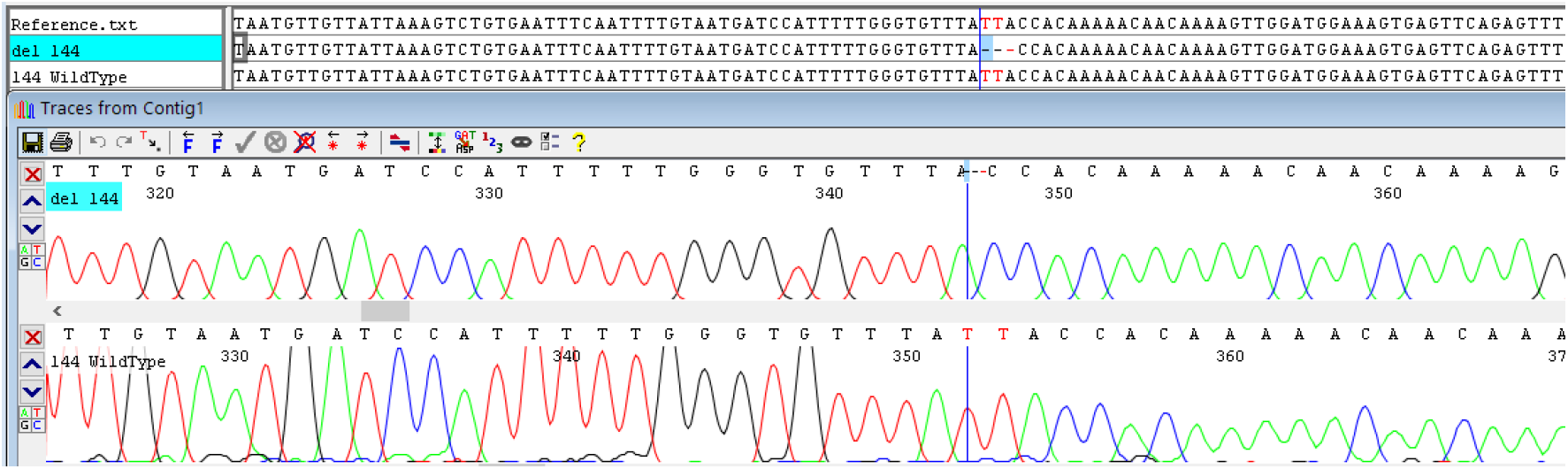
Chromatogram showing del 144 mutation of SARS-CoV-2 spike protein. The WildType chromatogram without codon 144 deletion is shown for comparison.

**Figure 5.**
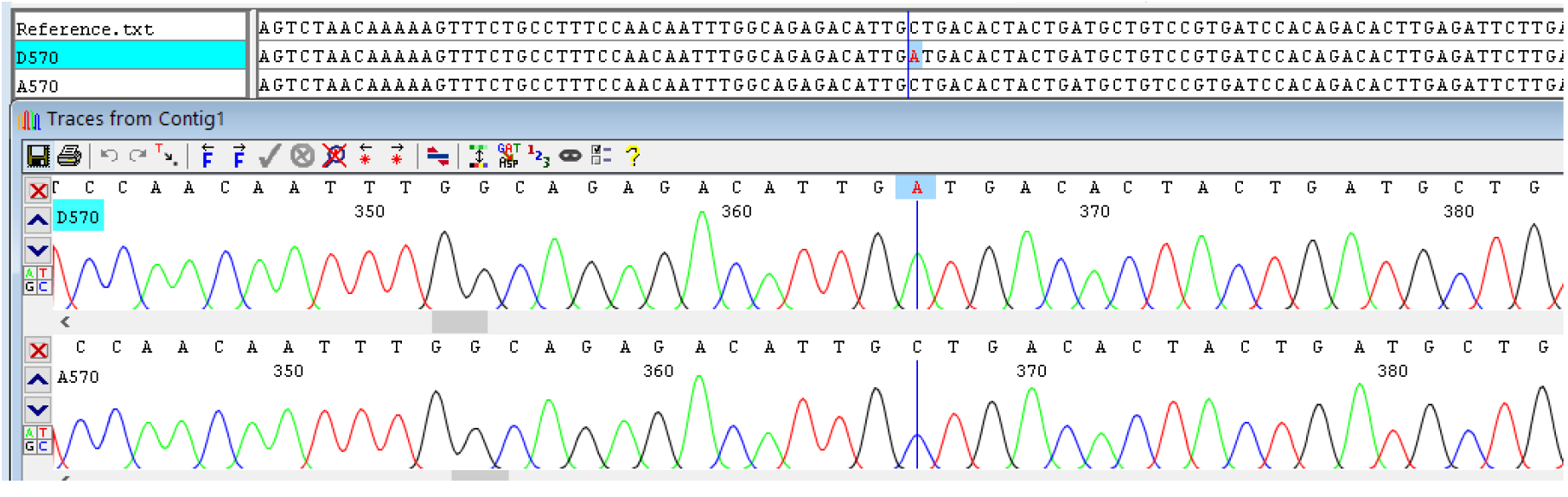
Chromatogram showing A570D (Ala570Asp) mutation of SARS-CoV-2 spike protein. The GCT>GAT conversion at codon 570 is highlighted.

**Figure 6.**
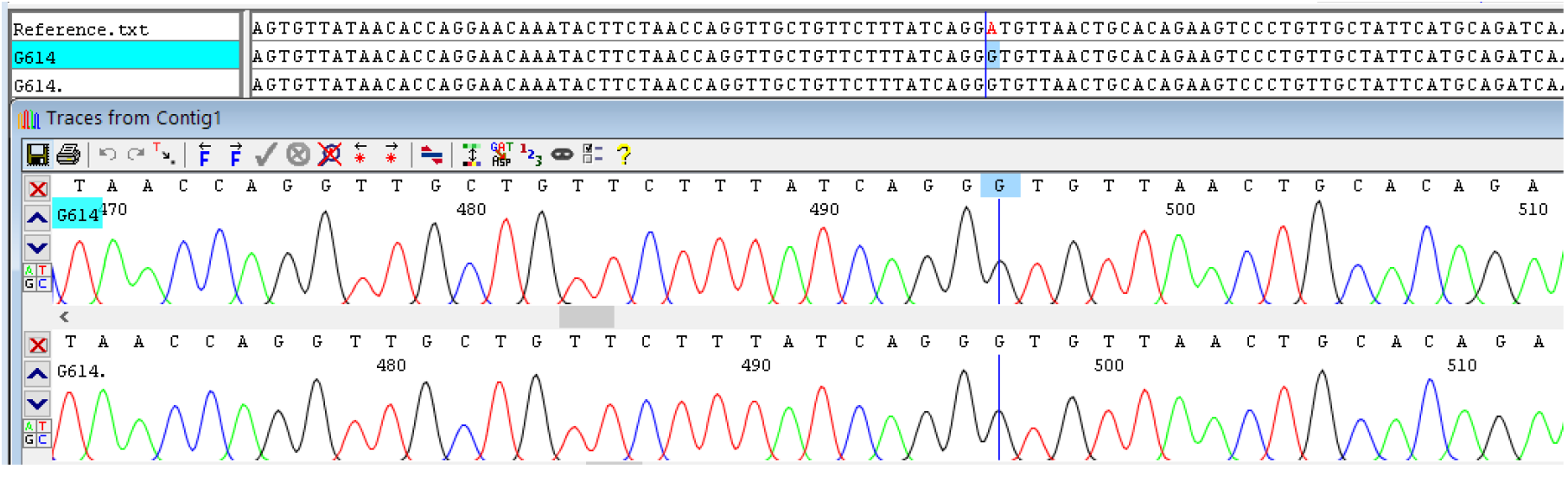
Chromatogram showing D614G (Asp614Gly) mutation of SARS-CoV-2 spike protein. The GAT>GGT conversion at codon 614 is highlighted. The D614 strain is no longer available.

**Figure 7.**
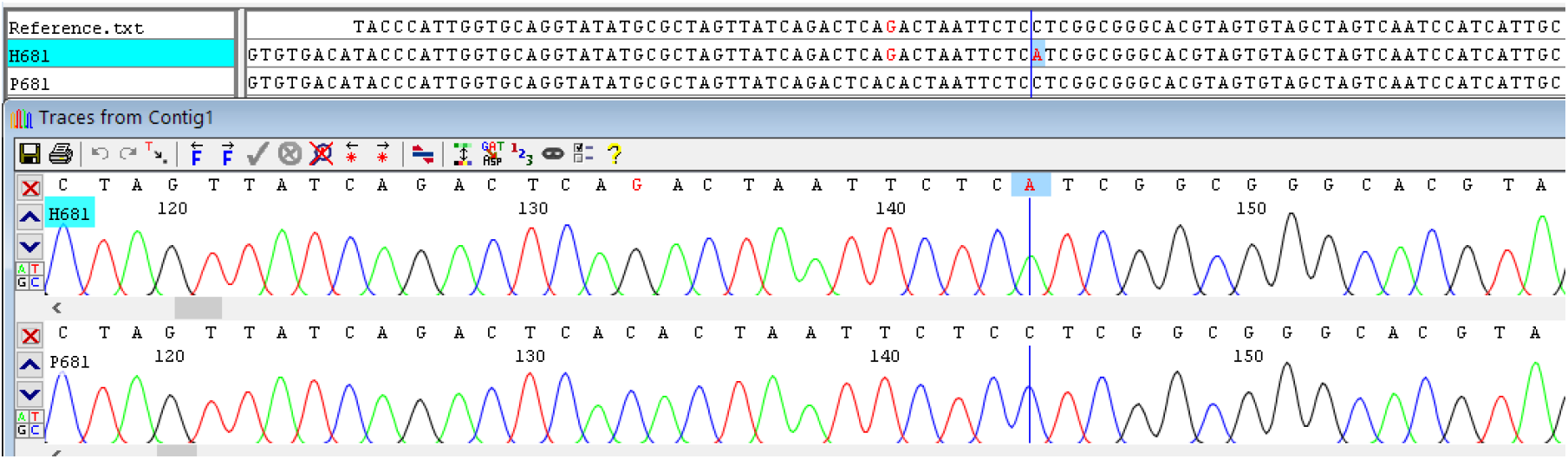
Chromatogram showing P681H (Pro681His) mutation of SARS-CoV-2 spike protein. The CCT>CAT conversion at codon 681 is highlighted.

**Figure 8.**
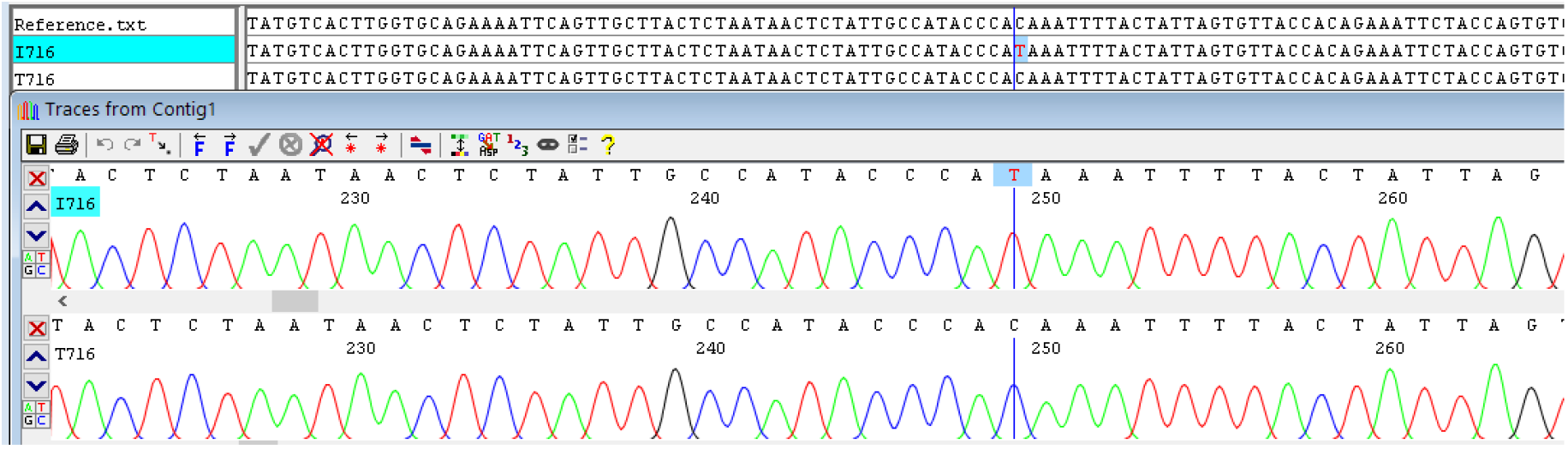
Chromatogram showing T716I (Thr716Ile) mutation of SARS-CoV-2 spike protein. The ACA>ATA conversion at codon 716 is highlighted.

**Figure 9.**
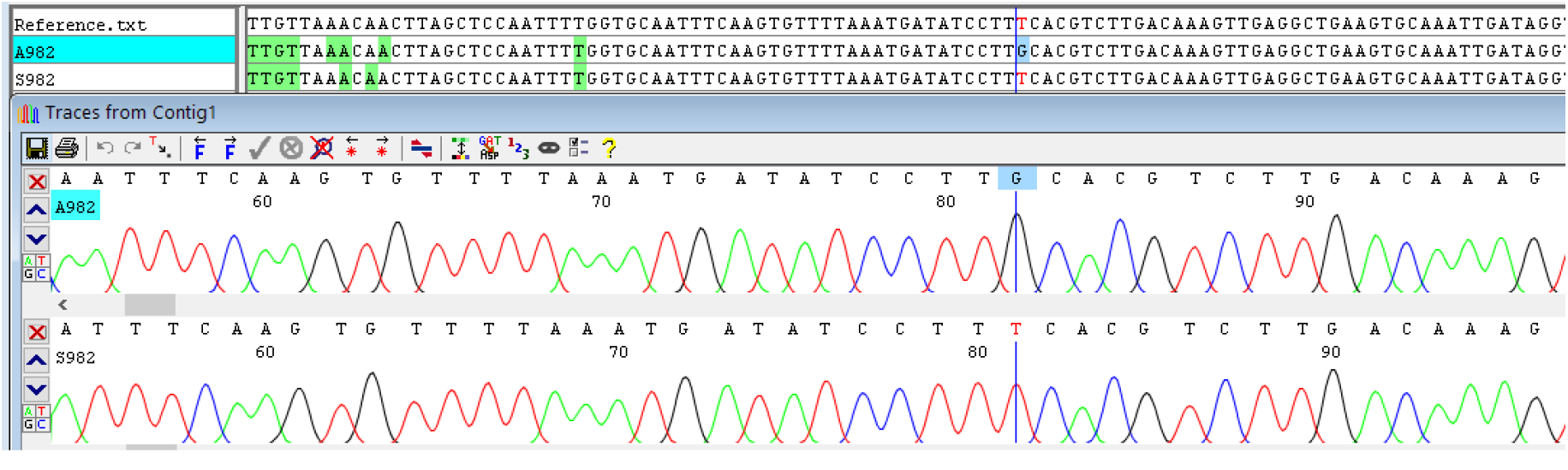
Chromatogram showing S982A (Ser982Ala) mutation of SARS-CoV-2 spike protein. The TCA>GCA conversion at codon 982 is highlighted.

**Figure 10.**
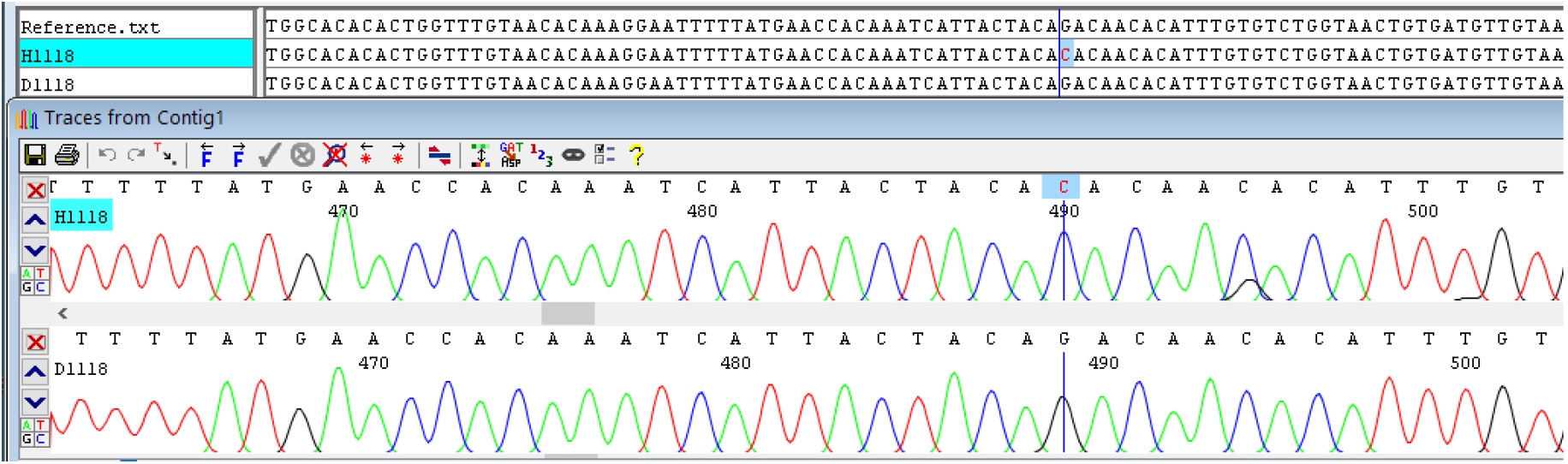
Chromatogram showing D1118H (Asp1118His) mutation of SARS-CoV-2 spike protein. The GAC>CAC conversion at codon 1118 is highlighted.

**Figure 11.**
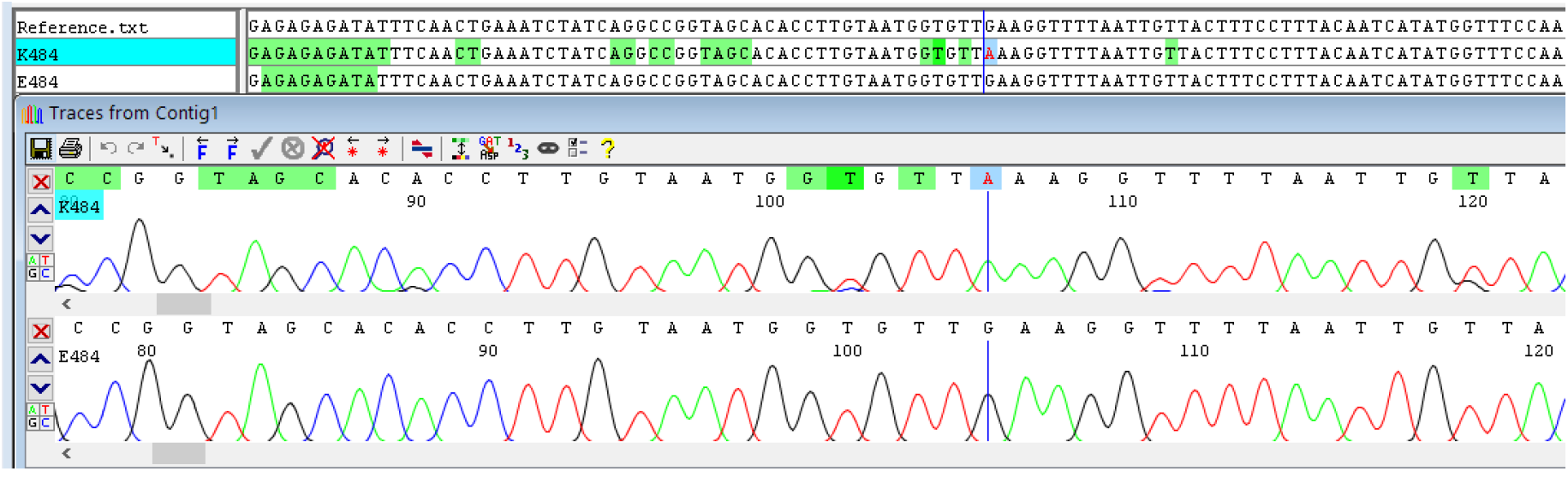
Chromatogram showing E484K (Glu484Lys) mutation of SARS-CoV-2 spike protein. The GAA>AAA conversion at codon 484 is highlighted.

## Discussion

The primer N501Y-1 specifically binds to the mutated Y501 strain but not to the wild type N501 strain. The probe N501Y-3 helps with the detection. The fluorophore at 5’ end of the probe can be modified in case of multiplexing along with the existing covid RT-PCR kits. In this study we have demonstrated single step cDNA conversion and amplification by traditional PCR for Sanger sequencing of targeted mutations.

Among the 12 samples of patients who tested positive for the UK Variant, Travel history –

Only 4 among them had a history of travel in the last 2 weeks-3 had just returned from Dubai 4 days before the date of sample collection and 1 had a history of returning from Maharashtra a week before the date of sample collection. None of the others had any travel history or any known history of coming in contact with someone who had recently travelled.

Clinical history –

7 out of 12 of the patients testing positive for the UK variant had only developed mild to moderate symptoms including myalgias, fatigue, mild fever, sore throat, headaches and loose stools. 5 of them had additional co-morbidities like Diabetes, Hypertension and Obesity and required hospitalization with supplemental Oxygen and were on Remdesivir, anti coagulants and other supportive medications and among these, 2 were on Non-invasive ventilation. Eventually, all of the 12 patients recovered well over a course of 3 weeks. Constant continued surveillance for newer variants of the SARS-Cov-2 and observing its clinical implications including any Vaccine-resistant strains is hence much needed to fight and eventually put an end to this pandemic.

## Data Availability

The authors confirm that the data supporting the findings of this study are available within the article. Additional info can be provided by Corresponding author upon request.

